# Association of Vascular Risk Factors with the Risk of Parkinson’s Disease among Stroke Survivors—A Nationwide Population-Based Study

**DOI:** 10.1101/2024.12.06.24318632

**Authors:** Seo Yeon Yoon, Hyun Im Moon, Jin Hyung Jung, Sang Chul Lee, Kyungdo Han, Yong Wook Kim

## Abstract

**Background:** Stroke survivors usually have many vascular risk factors (VRFs), and recent studies have reported the association of VRFs with cognitive decline among stroke survivors, which are also associated with PD risk. Therefore, we aimed to investigate how VRFs influenced the risk of PD occurrence in individuals with stroke.

**Methods:** The Korean National Health Insurance Service database was used. We selected individuals who experienced new-onset stroke (ICD-10: I60-I64) between 2008 and 2016 and followed up until 2019. PD was diagnosed according to ICD-10 code G20 and rare and intractable disease registration code V124. Lifestyle factors were collected using self-reported questionnaires, and comorbidities were defined based on ICD-10 codes, medication, and laboratory and anthropometric findings. Cox proportional hazard analyses were conducted to evaluate the association of VRFs with PD incidence among stroke survivors.

**Results:** Among 284,799 stroke survivors, 3,274 cases of PD were reported over a median follow-up period of 5.04 years. In individuals with a history of stroke, smoking (former smoking, hazard ratio (HR) = 0.80, 95% confidence interval (CI), 0.72–0.88; current smoking, HR = 0.66, 95% CI, 0.58–0.76) and alcohol consumption (moderate drinking, HR = 0.83, 95% CI, 0.75–0.91; heavy drinking, HR = 0.74, 95% CI, 0.59–0.94) were associated with a decreased risk of PD. Physically active individuals who had a stroke showed a reduced trend for PD occurrence (*P* = 0.0508). As for the association between comorbidities and the PD risk, the PD risk increased among stroke survivors with diabetes mellitus (P for trend <.001), particularly in those with a duration of diabetes mellitus of ≥5 years (HR = 1.38, 95% CI, 1.25–1.52).

**Conclusions:** Our results suggest that VRFs are associated with PD occurrence among stroke survivors. Understanding and managing VRFs in relation to PD risk is needed for disease management after stroke.

## Introduction

Parkinson’s disease (PD) is the most common and fastest-growing movement disorder, affecting approximately 7 million individuals globally. With the increasing life expectancy of the overall population, this number is estimated to double by 2040. ^1^ The clinical emergence of PD likely involves various risk factors, including both genetic and environmental factors, that exert effects over years before the clinical onset of PD. However, the exact etiology of PD remains unclear. ^2,3^

Stroke is a common neurological disease in older people and the leading cause of long-term physical and cognitive disability. Managing vascular risk factors (VRFs) such as hypertension (HTN), diabetes mellitus (DM), and dyslipidemia is crucial for the primary prevention of stroke and even for secondary prevention of recurrent stroke.^4^ High blood pressure (BP), glucose, and low-density lipoprotein cholesterol levels have also been associated with cognitive decline and dementia,^5^ and a recent study has suggested that glucose levels were associated with faster cognitive decline among stroke survivors.^6^ VRFs might accelerate cognitive decline through cerebral microvascular injury, oxidative stress, inflammation, and neurodegeneration. ^7–9^ These mechanisms have also been identified as potential contributors to the pathophysiology of PD in previous studies. ^10^

There have been several studies on the association between PD and stroke. Previous studies have examined the risk of stroke in patients with PD with inconsistent results. Some studies have shown that the risk of stroke is significantly increased after a previous PD event, ^11–13^ others have shown a reduced risk of stroke during life in patients with PD, ^14^ and some have not found a relationship between stroke and PD at all. ^15^. However, the risk of PD among stroke survivors has been scarcely investigated. ^16^ A recent meta-analysis has reported that PD and stroke may have a common pathogenesis and may share preventive treatment measures. ^17^ Ischemia in the brain can lead to nerve cell damage and death, cerebral infarction, neuroinflammatory reactions, blood-brain barrier damage, oxidative stress, and cell apoptosis and necrosis. These mechanisms can lead to dopamine neuron damage and play an important role in the development and progression of PD. ^2,18,19^

VRFs have mainly been investigated with a focus on secondary prevention in stroke survivors. Because of the shared mechanisms between PD and stroke, identifying and managing PD-related VRFs could influence the risk of PD occurrence among stroke survivors. Therefore, we aimed to investigate how various VRFs among stroke survivors are related to the occurrence of PD. We also performed subgroup analysis by stroke type and sex to evaluate whether the association between VRFs and the risk of PD remained consistent among stroke survivors.

## Methods

### Data source

The Korean National Health Insurance Service (NHIS) is a mandatory health insurance coverage provided by the government to all Korean populations. Each medical institution electronically submits all medical facility utilization data to the NHIS for reimbursement purposes, and information on healthcare utilization is centralized in the NHIS database. The data contains a unique anonymous number for each individual and summarizes the demographic factors, type of insurance, diagnostic codes based on the International Classification of Diseases (ICD-10), procedures, medical costs claimed, and prescribed drugs. Additionally, the standardized National Health Screening Program (NHSP) is provided at least bi-annually to enrollees of the NHIS. The NHSP includes anthropometric measurements, assessment of BP, laboratory investigations, and a self-reported questionnaire on lifestyle factors, such as smoking, alcohol consumption, and physical activity.

### Standard Protocol Approvals, Registrations, and Patient Consents

This study was approved by the Institutional Review Board of Yonsei University Severance Hospital (4-2023-1428), which waived the need for informed consent.

### Study population

We included individuals who were newly diagnosed with stroke, which was defined as the primary diagnosis according to ICD-10 codes (I60-I64) between 2008 and 2016. We only included individuals who were admitted to the hospital with a diagnosis of stroke based on brain imaging evaluations, such as magnetic resonance imaging or computed tomography. We exclude individuals with a combined diagnosis of transient ischemic attack. Among the included participants, only those who attended NHSP within 2 years after receiving the stroke diagnosis were included. Individuals aged <40 years and those with missing data were excluded. We excluded those who had a prior diagnosis of PD before enrollment. We set a 1-year lag period for PD diagnosis; therefore, individuals with a diagnosis of PD within one year of enrollment were also excluded. Finally, 284,799 individuals who experienced a stroke were enrolled in the analysis and followed up until December 31, 2019 (Figure 1).

**Figure 1.**
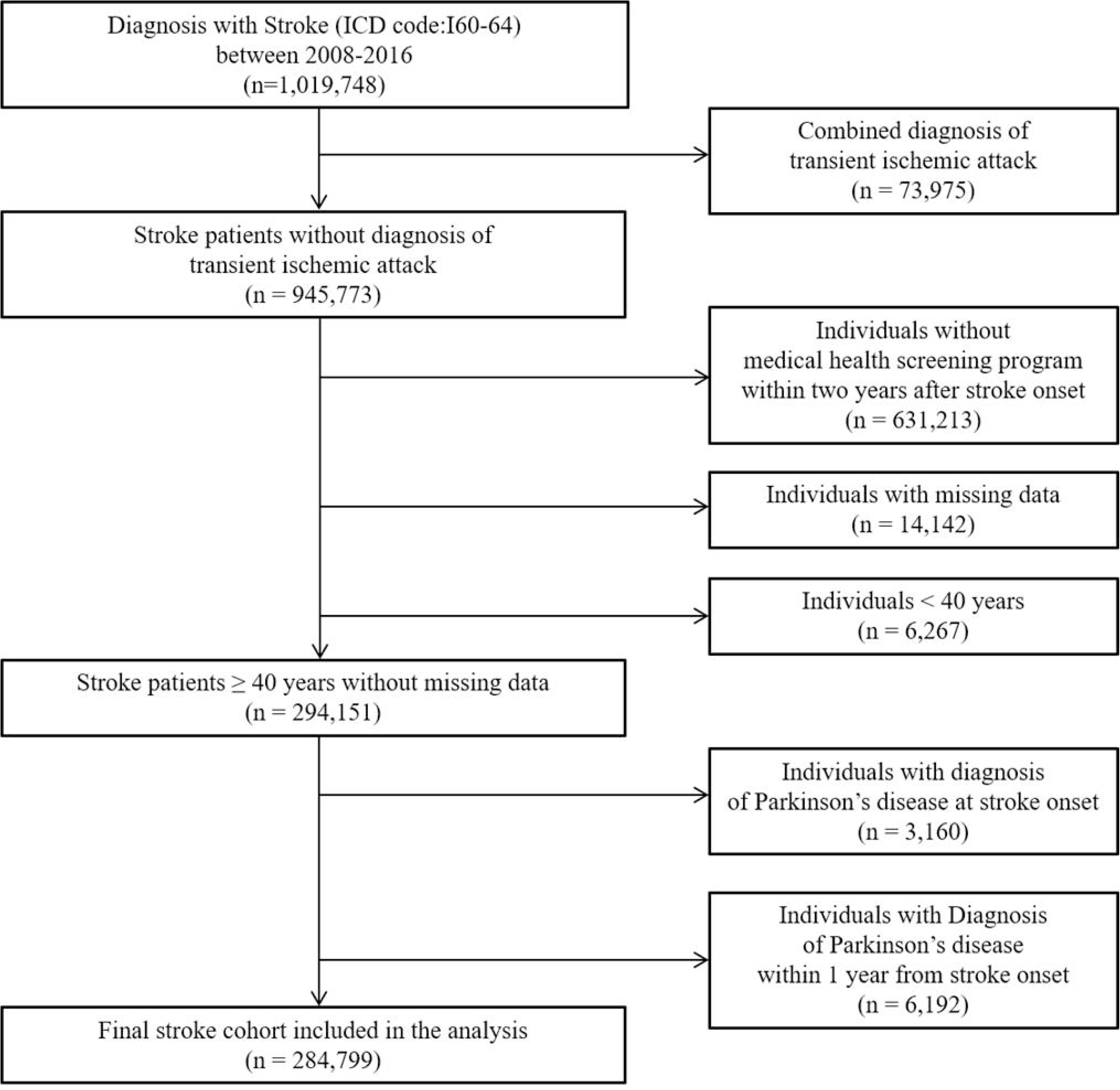
Flowchart for sample selection

### Definition of PD

The primary endpoint was newly diagnosed PD during the follow-up period. The Korean government has implemented a registration program for rare intractable diseases (RID), including PD, to assist patients with medical expenses since 2006. The diagnostic criteria for the V124 code are almost identical to the UK Parkinson’s Disease Society Brain Bank diagnostic criteria; therefore, the RID data are considered very reliable. Newly diagnosed PD was defined based on the ICD-10 code for PD (G20) and a registration code for PD (V124) in the RID program. In the sensitivity analysis, we excluded those with a combined diagnosis of atypical or secondary parkinsonism (ICD-10 codes: G21-G23) to ensure the study group consisted of individuals with homogenous PD.

### Other variables

Low income was defined as the bottom 25% according to the amount spent on insurance premiums as a proxy for income status. Height, weight, waist circumference, and BP (systolic and diastolic) were assessed. Body mass index (BMI) was calculated as the weight divided by height squared (kg/m^2^). Venous samples were drawn after an overnight fast to determine fasting plasma glucose and total cholesterol (TC). Detailed information on the health behaviors was obtained through standardized self-reported questionnaires. Participants were classified according to their smoking status as never, former, or current smokers. Alcohol consumption is categorized as none, mild to moderate (<30 g/d), or heavy (≥30 g/d). Physically active individuals were defined as vigorous-intensity exercise three or more times per week or moderate-intensity exercise five or more times per week. Baseline comorbidities (DM, HTN, and dyslipidemia) and the status of those comorbidities were identified based on a combination of ICD-10 codes, laboratory and anthropometric findings on NHSP, and data from the prescription database. DM was classified based on glycemic status as no DM (fasting glucose, 100 mg/dL), impaired fasting glucose (fasting glucose 100–125 mg/dL), incident DM (fasting glucose ≥ 126 mg/dL without DM medication), DM duration <5 years, and DM duration ≥5 years. HTN was classified based on BP as no HTN (SBP <120 mmHg and DBP <80 mmHg), pre-HTN (SBP 120–139 mmHg or DBP 80–89 mmHg), incident HTN (SBP ≥140 mmHg or DBP ≥90 mmHg without HTN medication), controlled HTN (SBP <140 mmHg and DBP <90 mmHg with HTN medication), and uncontrolled HTN (SBP ≥140 mmHg or DBP ≥90mmHg with HTN medication). Dyslipidemia was classified based on TC level as no dyslipidemia (TC <200 mg/dL), pre-dyslipidemia (TC 200–239 mg/dL), incident dyslipidemia (TC ≥240 mg/dL without dyslipidemia medication), controlled dyslipidemia (TC <240 mg/dL with dyslipidemia medication), and uncontrolled dyslipidemia (TC ≥240 mg/dL with dyslipidemia medication).

### Statistical analysis

Baseline characteristics of individuals who had a stroke are presented as the mean ± standard deviation for continuous variables or as the number (percentage) for categorical variables. Values were compared using an independent t-test or chi-square test for continuous and categorical variables, respectively. The incidence of PD was calculated by dividing the number of events by 1000 person-years. We conducted Cox proportional hazards analyses to evaluate the association of various risk factors, including socioeconomic status, lifestyle factors, and comorbidities, with the incidence of PD in individuals who had a stroke and estimated the hazard ratio (HR) and 95% confidence interval (CI). Model 1 was unadjusted. Model 2 was adjusted for age, sex, income, lifestyle factors, BMI, and comorbidities. Model 3 was further adjusted for mental disorders and antiplatelet use. Sensitivity analysis was performed after the exclusion of the combined diagnosis of atypical or secondary parkinsonism from the PD group to ensure homogeneity. Stratified analyses according to sex and stroke type were also performed. All statistical analyses were performed using SAS 9.4 (SAS Institute Inc., Cary, NC, USA), and a two-sided *P* < 0.05 was considered statistically significant.

### Data availability

## Results

### Characteristics of the study population

Table 1 presents the characteristics of the 284,799 individuals who had a stroke according to the stroke type. Among individuals who had a stroke, 225,513 (79.2%), 46,007 (16.2%), and 13,729 (4.7%) were identified as ischemic, hemorrhagic, and unspecified stroke types, respectively. The ischemic stroke group had a higher proportion of men, and the mean age of stroke onset was approximately 6 years older than those of hemorrhagic stroke. Individuals who had ischemic stroke had a higher prevalence of obesity (BMI ≥25 kg/m^2^), DM, HTN, and dyslipidemia than those with hemorrhagic stroke.

**Table 1.**
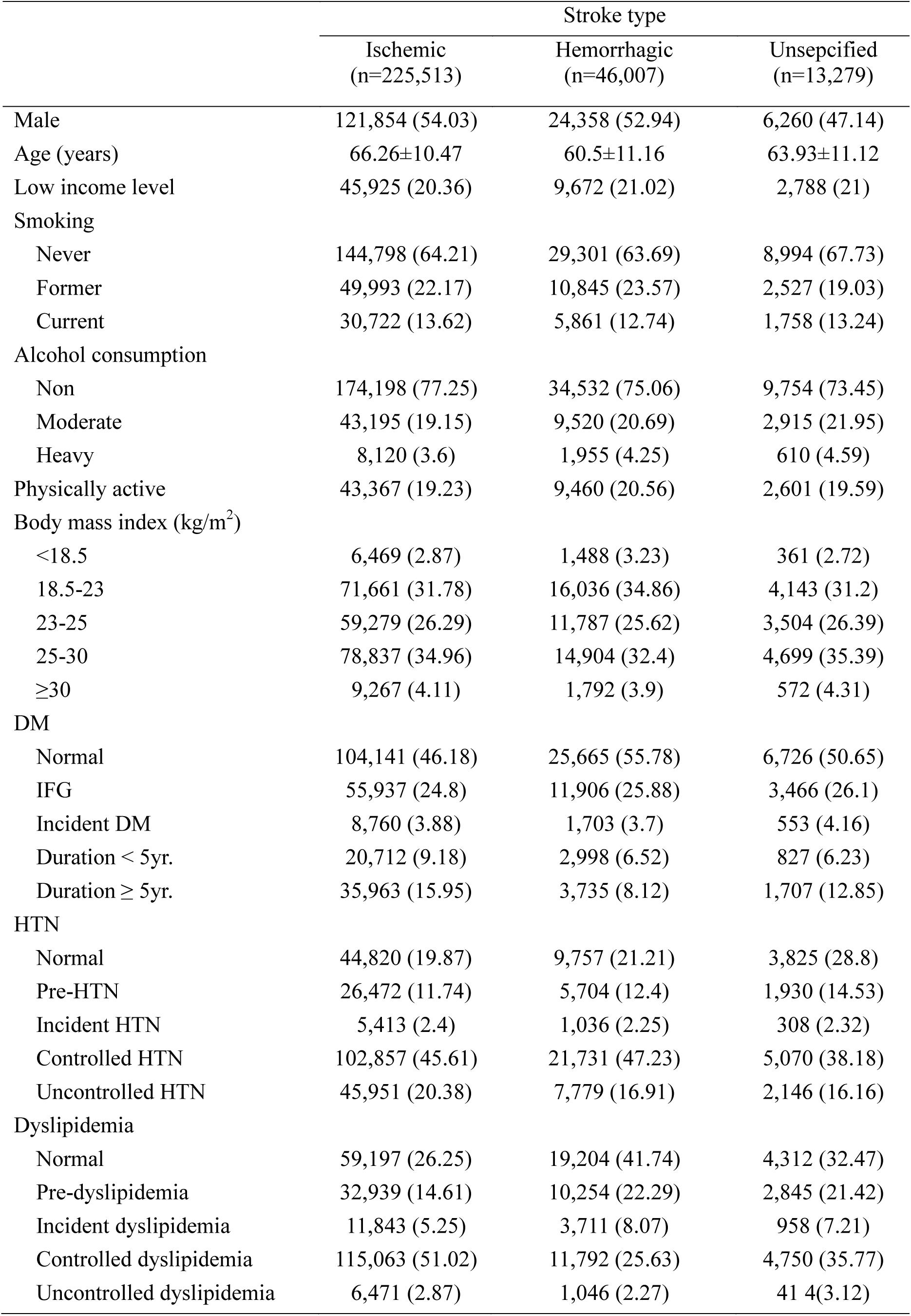
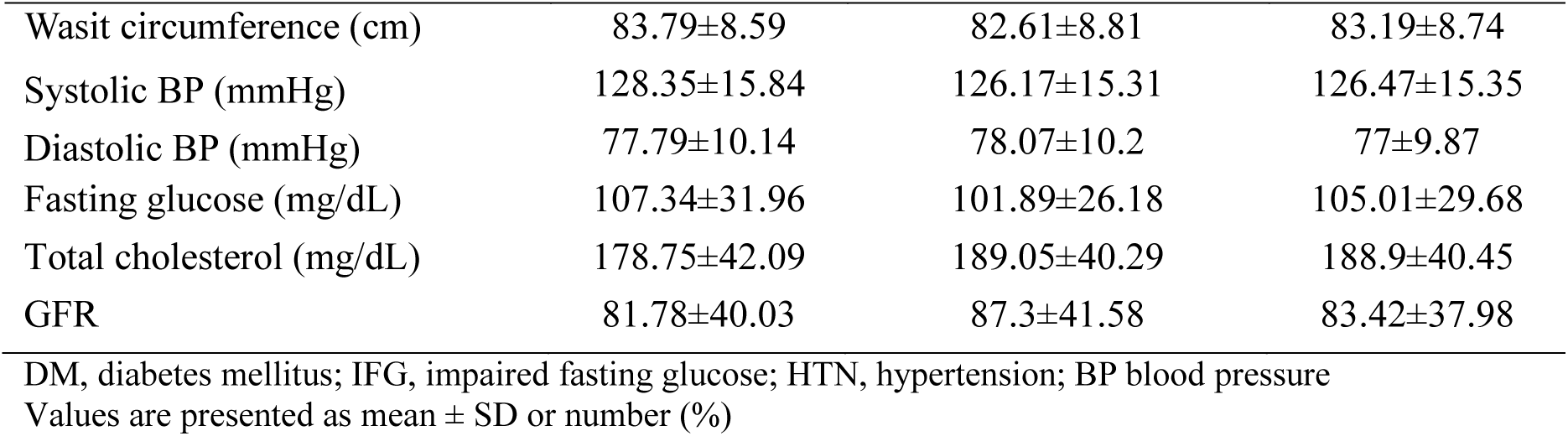
Characteristics of individuals with stroke.

|  | Stroke type |  |  |
| --- | --- | --- | --- |
|  | Ischemic<br>(n=225,513) | Hemorrhagic<br>(n=46,007) | Unsepcified<br>(n=13,279) |
| Male | 121,854 (54.03) | 24,358 (52.94) | 6,260 (47.14) |
| Age (years) | 66.26±10.47 | 60.5±11.16 | 63.93±11.12 |
| Low income level | 45,925 (20.36) | 9,672 (21.02) | 2,788 (21) |
| Smoking |  |  |  |
| Never | 144,798 (64.21) | 29,301 (63.69) | 8,994 (67.73) |
| Former | 49,993 (22.17) | 10,845 (23.57) | 2,527 (19.03) |
| Current | 30,722 (13.62) | 5,861 (12.74) | 1,758 (13.24) |
| Alcohol consumption |  |  |  |
| Non | 174,198 (77.25) | 34,532 (75.06) | 9,754 (73.45) |
| Moderate | 43,195 (19.15) | 9,520 (20.69) | 2,915 (21.95) |
| Heavy | 8,120 (3.6) | 1,955 (4.25) | 610 (4.59) |
| Physically active | 43,367 (19.23) | 9,460 (20.56) | 2,601 (19.59) |
| Body mass index (kg/m<sup>2</sup>) |  |  |  |
| <18.5 | 6,469 (2.87) | 1,488 (3.23) | 361 (2.72) |
| 18.5-23 | 71,661 (31.78) | 16,036 (34.86) | 4,143 (31.2) |
| 23-25 | 59,279 (26.29) | 11,787 (25.62) | 3,504 (26.39) |
| 25-30 | 78,837 (34.96) | 14,904 (32.4) | 4,699 (35.39) |
| ≥30 | 9,267 (4.11) | 1,792 (3.9) | 572 (4.31) |
| DM |  |  |  |
| Normal | 104,141 (46.18) | 25,665 (55.78) | 6,726 (50.65) |
| IFG | 55,937 (24.8) | 11,906 (25.88) | 3,466 (26.1) |
| Incident DM | 8,760 (3.88) | 1,703 (3.7) | 553 (4.16) |
| Duration < 5yr. | 20,712 (9.18) | 2,998 (6.52) | 827 (6.23) |
| Duration ≥ 5yr. | 35,963 (15.95) | 3,735 (8.12) | 1,707 (12.85) |
| HTN |  |  |  |
| Normal | 44,820 (19.87) | 9,757 (21.21) | 3,825 (28.8) |
| Pre-HTN | 26,472 (11.74) | 5,704 (12.4) | 1,930 (14.53) |
| Incident HTN | 5,413 (2.4) | 1,036 (2.25) | 308 (2.32) |
| Controlled HTN | 102,857 (45.61) | 21,731 (47.23) | 5,070 (38.18) |
| Uncontrolled HTN | 45,951 (20.38) | 7,779 (16.91) | 2,146 (16.16) |
| Dyslipidemia |  |  |  |
| Normal | 59,197 (26.25) | 19,204 (41.74) | 4,312 (32.47) |
| Pre-dyslipidemia | 32,939 (14.61) | 10,254 (22.29) | 2,845 (21.42) |
| Incident dyslipidemia | 11,843 (5.25) | 3,711 (8.07) | 958 (7.21) |
| Controlled dyslipidemia | 115,063 (51.02) | 11,792 (25.63) | 4,750 (35.77) |
| Uncontrolled dyslipidemia | 6,471 (2.87) | 1,046 (2.27) | 414 (3.12) |
| Wasit circumference (cm) | 83.79±8.59 | 82.61±8.81 | 83.19±8.74 |
| Systolic BP (mmHg) | 128.35±15.84 | 126.17±15.31 | 126.47±15.35 |
| Diastolic BP (mmHg) | 77.79±10.14 | 78.07±10.2 | 77±9.87 |
| Fasting glucose (mg/dL) | 107.34±31.96 | 101.89±26.18 | 105.01±29.68 |
| Total cholesterol (mg/dL) | 178.75±42.09 | 189.05±40.29 | 188.9±40.45 |
| GFR | 81.78±40.03 | 87.3±41.58 | 83.42±37.98 |

DM, diabetes mellitus; IFG, impaired fasting glucose; HTN, hypertension; BP blood pressure Values are presented as mean ± SD or number (%)

### Risk factors for PD occurrence in individuals who had a stroke

Table 2 shows various risk factors for the risk of PD occurrence. During the follow-up period (median 5.04 years, interquartile range 2.96–7.28), 3,274 cases of PD (1.15%) occurred in individuals with stroke. Male sex (HR = 1.42, 95% CI, 1.30–1.54) and older age (HR = 3.28, 95% CI, 3.00–3.58) were significantly associated with an increased risk of PD. In individuals who had a stroke, former (HR = 0.80, 95% CI, 0.72–0.88) and current (HR = 0.66, 95% CI, 0.58–0.76) smoking showed a significant association with reduced risk of PD. Alcohol consumption was also associated with a decreased risk of PD in individuals who had a stroke (moderate drinking, HR = 0.83, 95% CI, 0.75–0.91; heavy drinking, HR = 0.74, 95% CI, 0.59–0.94). Physically active individuals who had a stroke showed a reduced trend for PD occurrence (HR = 0.91, 95% CI, 0.83-–.00, *P* = 0.0508). As for the association between comorbidities and PD risk, the risk of PD increased among stroke survivors with DM (P for trend <.001), particularly among stroke survivors with DM duration ≥5 years, the PD risk increased by 38% (HR = 1.38, 95% CI, 1.25–1.52). The PD risk was reduced in patients who had a stroke with dyslipidemia (P for trend = 0.0003), whereas there were no significant associations of BMI and HTN with the risk of PD in individuals who had a stroke (*P* >0.05)

**Table 2.**
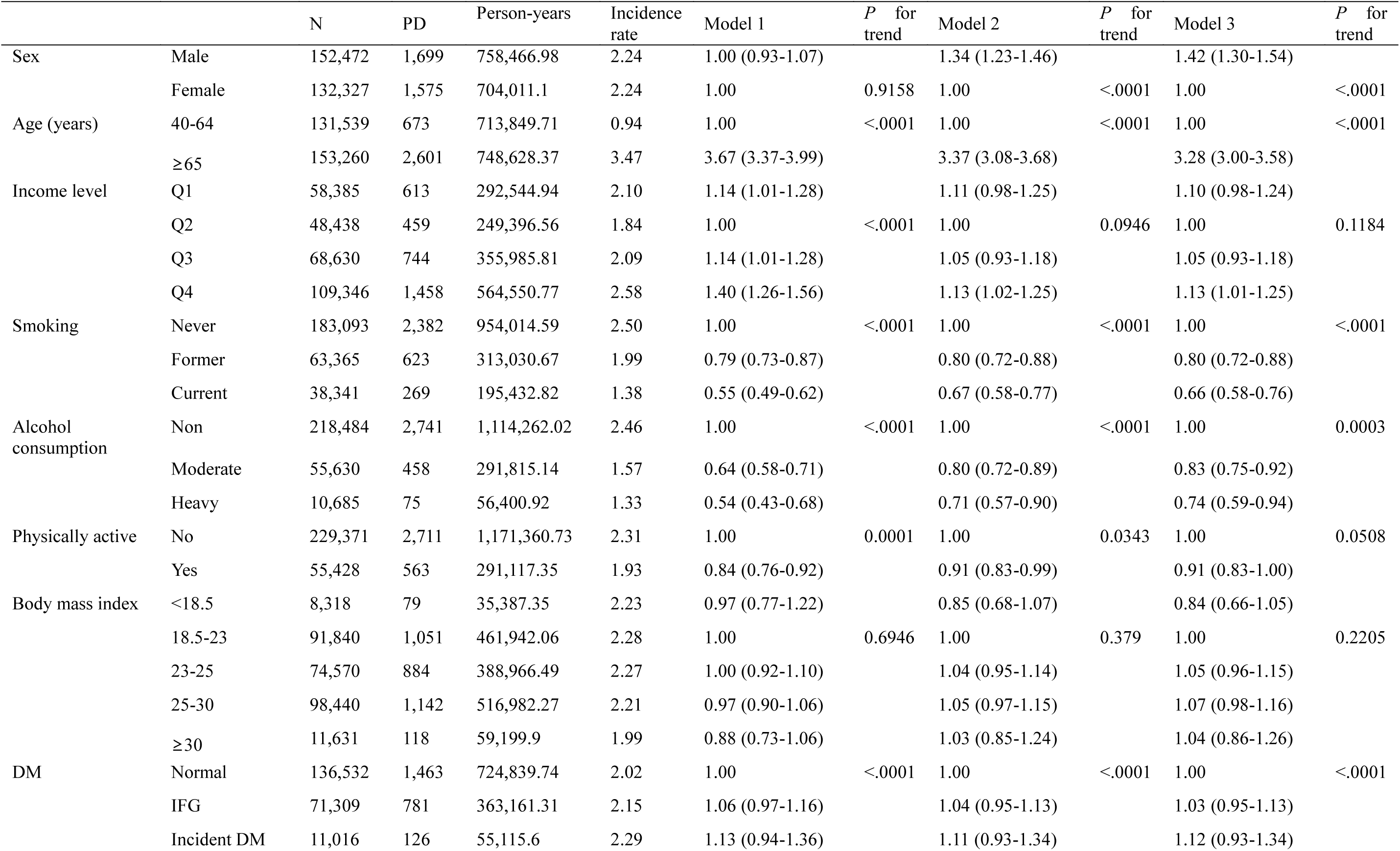
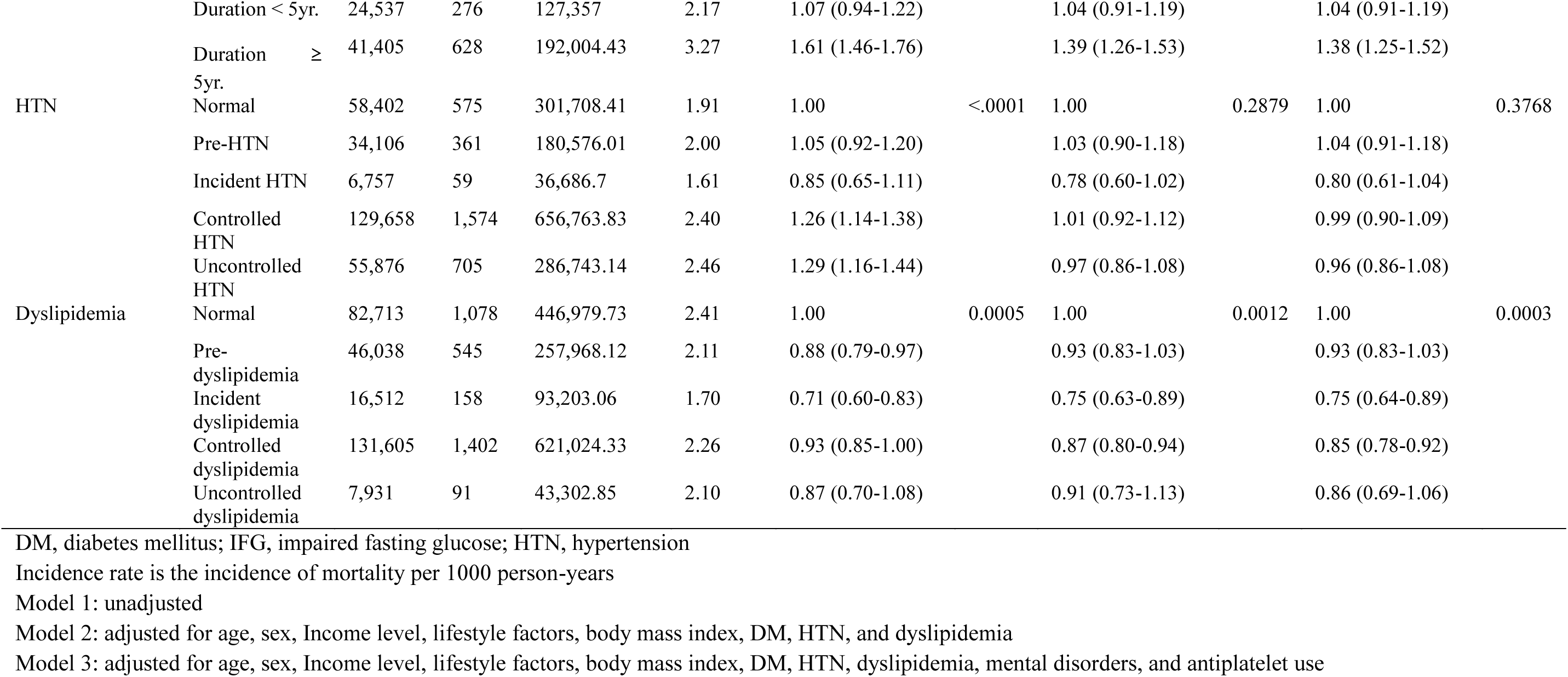
Cox Proportional Hazard Regression Analysis of the Parkinson’s Disease Risk in Individuals with Stroke.

When a combined diagnosis of secondary or atypical parkinsonism was excluded from the diagnosis of PD for the sensitivity analysis, the results showed almost similar findings to the main results (Supplementary Table 1).

### Subgroup analysis by sex and stroke type

Table 3 presents the association of various factors with PD risk in individuals who had a stroke stratified by sex. There were significant differences in the associations of smoking and dyslipidemia with PD risk by sex. Smoking (former, HR = 0.80, 95% CI, 0.78–0.89; current, HR = 0.66, 95% CI, 0.57–0.77, P for trend <.0001) and dyslipidemia (incident dyslipidemia, HR = 0.64, 95% CI, 0.47–0.86; controlled dyslipidemia, HR = 0.82, 95% CI, 0.73–0.92, P for trend = 0.0008) were significantly associated with a reduced PD risk only in males. DM showed a significant association with PD risk in both males (P for trend = 0.0111) and females (P for trend <.0001).

**Table 3.** Subgroup analysis of the Parkinson’s Disease Risk in Individuals with Stroke.

|  |  | Male | *P* for trend | Female | *P* for trend |
| --- | --- | --- | --- | --- | --- |
| Age | 40-64 | 1.00 | <.0001 | 1.00 | <.0001 |
|  | ≥65 | 3.27 (2.90-3.68) |  | 3.20 (2.80-3.67) |  |
| Income level | Q1 | 1.06 (0.89-1.25) |  | 1.15 (0.97-1.37) |  |
|  | Q2 | 1.00 | 0.071 | 1.00 | 0.4566 |
|  | Q3 | 1.02 (0.87-1.20) |  | 1.08 (0.92-1.28) |  |
|  | Q4 | 1.16 (1.01-1.35) |  | 1.08 (0.93-1.26) |  |
| Smoking | Never | 1.00 | <.0001 | 1.00 | 0.0551 |
|  | Former | 0.80 (0.78-0.89) |  | 0.66 (0.39-1.09) |  |
|  | Current | 0.66 (0.57-0.77) |  | 0.67 (0.43-1.04) |  |
| Alcohol consumption | Non | 1.00 | 0.0184 | 1.00 | 0.0001 |
|  | Moderate | 0.90 (0.80-1.01) |  | 0.51 (0.37-0.70) |  |
|  | Heavy | 0.75 (0.59-0.95) |  | 1.20 (0.39-3.76) |  |
| Physically active | No | 1.00 | 0.2248 | 1.00 | 0.1108 |
|  | Yes | 0.93 (0.83-1.05) |  | 0.88 (0.76-1.03) |  |
| Body mass index (kg/m<sup>2</sup>) | <18.5 | 0.93 (0.68-1.28) |  | 0.75 (0.53-1.04) |  |
|  | 18.5-23 | 1.00 | 0.8218 | 1.00 | 0.3095 |
|  | 23-25 | 1.04 (0.92-1.18) |  | 1.06 (0.92-1.20) |  |
|  | 25-30 | 1.07 (0.95-1.20) |  | 1.06 (0.94-1.20) |  |
|  | ≥30 | 1.02 (0.73-1.41) |  | 1.04 (0.82-1.32) |  |
| DM | Normal | 1.00 | 0.0111 | 1.00 | <.0001 |
|  | IFG | 0.98 (0.87-1.11) |  | 1.09 (0.96-1.24) |  |
|  | Incident DM | 1.10 (0.86-1.39) |  | 1.13 (0.85-1.49) |  |
|  | Duration < 5yr. | 0.92 (0.77-1.11) |  | 1.18 (0.98-1.43) |  |
|  | Duration ≥ 5yr. | 1.23 (1.08-1.41) |  | 1.55 (1.35-1.78) |  |
| HTN | Normal | 1.00 | 0.455 | 1.00 | 0.5987 |
|  | Pre-HTN | 1.01 (0.84-1.20) |  | 1.07 (0.88-1.30) |  |
|  | Incident HTN | 0.73 (0.50-1.06) |  | 0.87 (0.60-1.27) |  |
|  | Controlled HTN | 1.03 (0.90-1.18) |  | 0.95 (0.82-1.10) |  |
|  | Uncontrolled HTN | 0.98 (0.83-1.15) |  | 0.93 (0.79-1.10) |  |
| Dyslipidemia | Normal | 1.00 | 0.0008 | 1.00 | 0.1507 |
|  | Pre-dyslipidemia | 0.91 (0.79-1.05) |  | 0.95 (0.82-1.11) |  |
|  | Incident dyslipidemia | 0.64 (0.47-0.86) |  | 0.83 (0.68-1.03) |  |
|  | Controlled dyslipidemia | 0.82 (0.73-0.92) |  | 0.89 (0.78-1.01) |  |
|  | Uncontrolled dyslipidemia | 1.06 (0.75-1.50) |  | 0.78 (0.59-1.02) |  |

DM, diabetes mellitus; IFG, impaired fasting glucose; HTN, hypertension
Adjusted for age, income level, lifestyle factors, body mass index, DM, HTN, dyslipidemia, mental disorders, and antiplatelet use

The association of various factors with PD risk in individuals who had a stroke stratified by stroke type is presented in Figure 2. Overall, smoking (P for trend <.0001), alcohol consumption (P for trend = 0.0007), dyslipidemia (P for trend <.0001), and DM (P for trend <.0001) showed more significant association with PD in individuals with ischemic stroke than hemorrhagic stroke.

**Figure 2.**
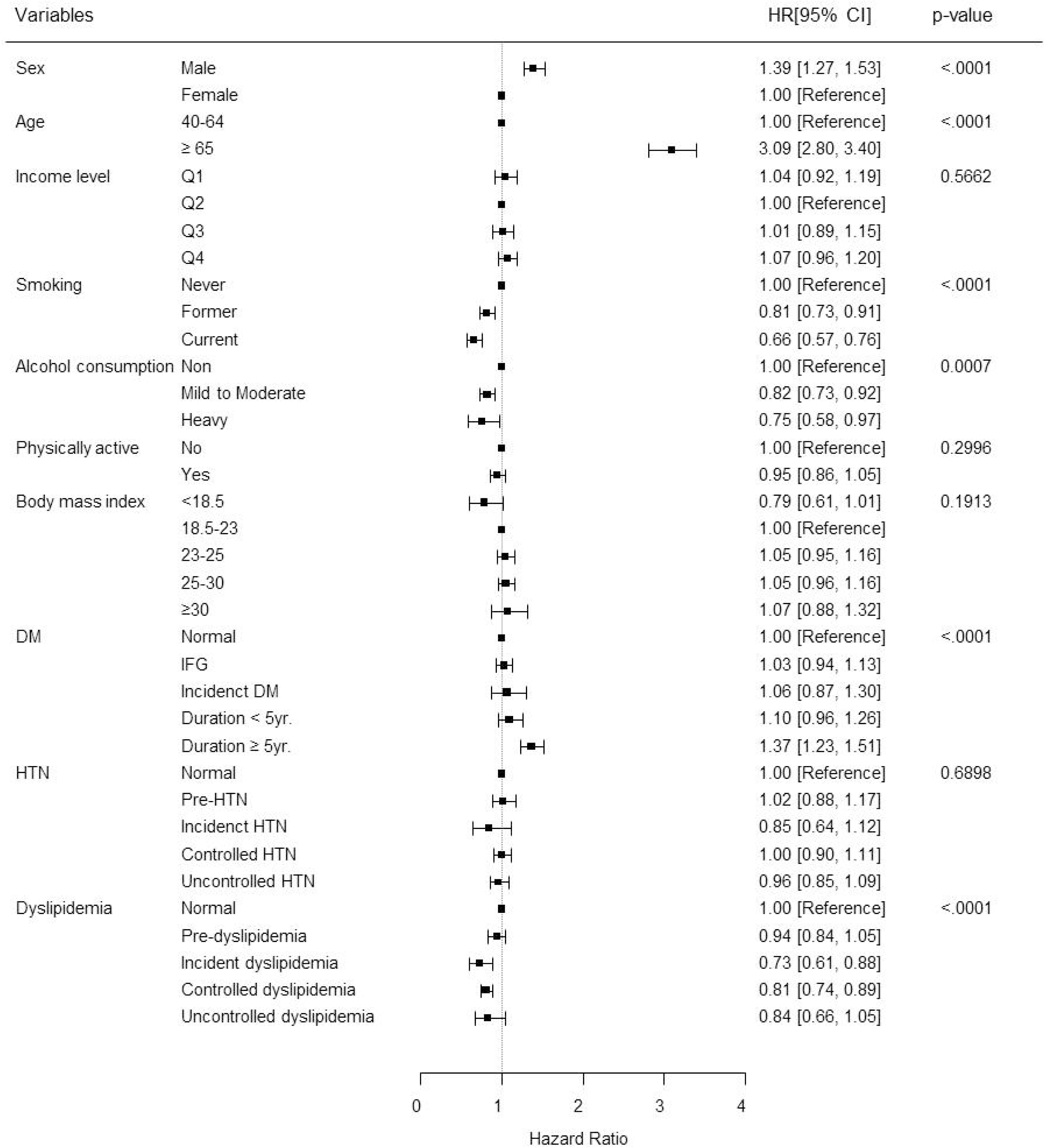
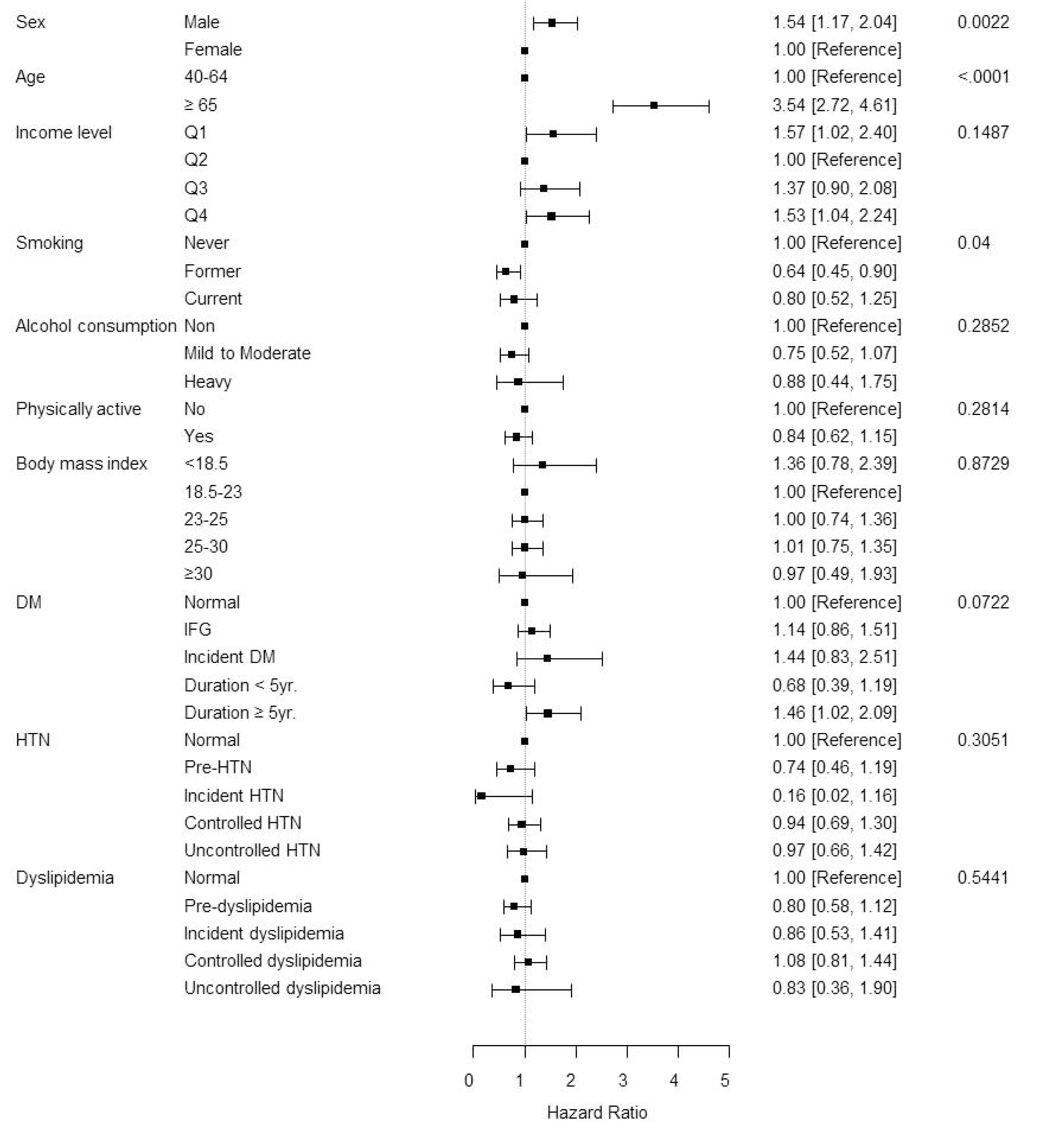
Subgroup analysis of the Parkinson’s disease risk in individuals with stroke according to stroke type. (a) ischemic stroke and (b) hemorrhagic stroke

## Discussion

In the present study, we analyzed 284,799 individuals who had a stroke to evaluate PD development according to various VRFs using nationwide population-based cohort data comprising the entire Korean population. Male sex, older age, and the presence of DM were significantly associated with increased risk of PD among stroke survivors, whereas the presence of dyslipidemia showed an association with reduced risk of PD. As for lifestyle factors, smoking and alcohol consumption reduced PD risk, and the physically active lifestyle had some trend for reduced PD occurrence.

The absolute number of people affected by stroke (i.e., who die from or remain disabled by stroke) has almost doubled during the past three decades, with >86% of the stroke burden in low-income and middle-income countries. ^20^ Therefore, patients who had a stroke often live with restrictions on cognition and physical function as sequelae. VRFs have been considered important for secondary prevention in stroke survivors, and recently, a correlation between cognitive decline after stroke and VRFs has been reported. ^6,21^ Patients who had a stroke often have poor physical dysfunction, and VRFs may cause motor dysfunction, such as vascular parkinsonism, worsening physical function in patients who had a stroke. PD is the second most common neurodegenerative disease that occurs frequently in old age, and studies on the association between VRFs and PD occurrence have been published recently. ^15,22,23^ Given that PD occurrence in patients who had a stroke can worsen their functional level, making long-term prognosis worse, our study aimed to analyze the relationship between VRFs and PD occurrence. Our findings suggest that various VRFs could influence the risk of PD occurrence among stroke survivors, which raises awareness of the importance of managing VRFs even for physical function among stroke survivors.

In our results, old age and male sex were significant risk factors for PD occurrence among stroke survivors, which is in line with previous studies conducted in the general population. ^24,25^ As for the association between comorbidities and PD risk, the PD risk significantly increased in patients who had a stroke with DM, particularly in the DM duration ≥5 years group. Recently, many studies have reported a positive relationship between DM and PD risk, and it has been reported that even the glycemic status is associated with PD risk. ^22,23,26^ As for the possible mechanisms between the two diseases, circulating insulin could have a neuroprotective role, whereas insulin resistance could affect pathways important in PD pathogenesis, such as mitochondrial dysfunction, neuroinflammation, and synaptic plasticity. ^23^ In terms of dyslipidemia, the PD risk was reduced in patients who had a stroke with dyslipidemia in this study. The association between dyslipidemia and PD remains controversial. Previous studies have suggested that statin use lowers the incidence of PD, and anti-inflammatory and neuroprotective effects have been suggested as the underlying mechanism. ^27,28^ However, there have also been many studies on the inverse association between PD and lipid levels. High cholesterol levels have also been reported to reduce the incidence of PD in a cohort without statin use. ^29,30^ In our results, there were no significant associations of BMI and HTN with the risk of PD in individuals who had a stroke. There have been fewer studies published focusing on the effect of a previous diagnosis of HTN on the risk of PD, and some studies yield reduced risk of PD, whereas other studies showed no effect. ^31–33^ Additionally, a recent meta-analysis has shown that patients with PD had a significantly lower BMI than controls, and some studies have shown an inverse association between BMI and PD. ^34^ Among the VRFs mentioned above, DM and dyslipidemia are thought to have a strong relationship with PD occurrence. A previous report has proposed a more specific interaction between VRFs and PD, which involves accelerating the neurodegenerative process, particularly in the presence of DM. ^35^

In our study, former and current smoking showed a significant association with a reduced risk of PD. There seems to be some dilemma with this result. Patients who had a stroke should quit smoking to prevent recurrent strokes, so non-smoking is usually recommended in those patients. However, regarding the association between smoking and PD, there have been many studies supporting the reduced PD risk among smokers, which is in line with our findings. A previous meta-analysis has reported that smokers have a lower risk of PD. ^36^ Another epidemiologic study has also reported that ever having smoked cigarettes was associated with a reduced risk of PD. ^37^ Alcohol consumption was also associated with a decreased risk of PD in individuals who had a stroke. There are still controversies regarding the effects of alcohol consumption on disease, not only in PD but also in stroke. Studies have shown that moderate drinking can reduce the risk of ischemic stroke by increasing high-density lipoprotein cholesterol and suppress blood clotting. However, excessive drinking can increase the risk of hemorrhagic and ischemic strokes. Alcohol abuse can worsen stroke risk factors, such as high BP, heart arrhythmia, and especially atrial fibrillation. ^38,39^ Likewise, the relationship between drinking and developing PD is unclear, and research results are inconsistent. Some studies have suggested that moderate drinking may slightly reduce the risk of PD, but others show that no clear relationship is found or drinking does not significantly affect the risk of PD. ^36,37,40^ The possible mechanism of the neuroprotective effect of moderate drinking is that antioxidants contained in alcoholic beverages, such as polyphenols, can protect dopaminergic neurons. ^41^

Physically active individuals who had a stroke showed a reduced trend for PD occurrence, although it is not statistically significant. Physical activity or exercise is recommended for stroke survivors to reduce the risk of recurrent stroke, and there has been evidence that physical activity reduces PD risk. ^42^ Stroke survivors often have some functional limitations, and there can be a possibility that moderate or vigorous physical activity would not be performed efficiently, potentially impacting our results.

In the subgroup analysis by sex, smoking and dyslipidemia were significantly associated with a reduced PD risk only in males. In the case of smoking, women’s smoking rate is extremely low in Korea, so it may have been difficult to draw statistically meaningful results. As for dyslipidemia, the complex interaction between sex and lipid metabolism in relation to PD is unclear. However, the modulatory effect of sex hormones such as estrogen and androgen on lipoprotein metabolism should be taken into account. ^43^ Given that there are many reports showing that the occurrence and clinical characteristics of PD differ according to sex, more studies are needed to confirm the characteristics of men and women. In subgroup analysis by stroke type, smoking, alcohol consumption, dyslipidemia, and DM showed a more significant association with PD in ischemic stroke than in hemorrhagic stroke. It might be originated from the characteristics of the population. In our study population, the ischemic stroke group had a higher prevalence of obesity, DM, HTN, and dyslipidemia than hemorrhagic stroke. Based on our results, VRFs need more attention and should be controlled properly in relation to PD risk, especially for ischemic stroke survivors.

This study has some limitations. First, we used claims-based data, and misclassification of PD and stroke would be considered. We attempted to use the strict operational definition of PD based on the ICD-code and rare and intractable disease registration code and also performed a sensitivity analysis after the exclusion of combined diagnosis of atypical or secondary parkinsonism from the PD group. For stroke, we only included individuals who were admitted to the hospital with a diagnosis of stroke based on brain imaging evaluations.

Second, although we tried to include many variables associated with PD occurrence, dietary habits, such as dietary food or caffeine consumption, were unavailable and were not included in the analysis. Finally, this study only included the Korean population, and caution should be needed when extrapolating these results to other ethnicities.

## Conclusion

Since patients who had a stroke have more VRFs than the general population, lifestyle interventions, including attention to controlling VRFs, especially DM, should be encouraged as part of a comprehensive approach in the management of patients with a new diagnosis of stroke to reduce the PD risk.

## Non-standard Abbreviations and Acronyms

VRF: vascular risk factors
PD: Parkinson’s disease
HTN: hypertension
DM: diabetes mellitus
BP: blood pressure
NHIS: National Health Insurance Service
ICD: International Classification of Diseases
NHSP: National Health Screening Program
TC: total cholesterol
MBI: Modified Barthel Index
HR: hazard ratio
CI: confidence interval
SES: socioeconomic status

## Data Availability

yes

## Acknowledgments

Funding: This study was supported by the National Research Foundation of Korea grant funded by the Korean government (MSIT) (RS-2024-00337352).

## Declaration of interest

None.

## Notes

### Competing Interest Statement

The authors have declared no competing interest.

## References

1. Dorsey ER, Bloem BR. The Parkinson Pandemic-A Call to Action. JAMA Neurol. 2018;75:9–10. doi: 10.1001/jamaneurol.2017.3299

2. Saiki S. [The association of Parkinson’s disease pathogenesis with inflammation]. Rinsho Shinkeigaku. 2014;54:1125–1127. doi: 10.5692/clinicalneurol.54.1125

3. Dardiotis E, Xiromerisiou G, Hadjichristodoulou C, Tsatsakis AM, Wilks MF, Hadjigeorgiou GM. The interplay between environmental and genetic factors in Parkinson’s disease susceptibility: the evidence for pesticides. Toxicology. 2013;307:17–23. doi: 10.1016/j.tox.2012.12.016

4. Kleindorfer DO, Towfighi A, Chaturvedi S, Cockroft KM, Gutierrez J, Lombardi-Hill D, Kamel H, Kernan WN, Kittner SJ, Leira EC, et al. 2021 Guideline for the Prevention of Stroke in Patients With Stroke and Transient Ischemic Attack: A Guideline From the American Heart Association/American Stroke Association. Stroke. 2021;52:e364–e467. doi: 10.1161/str.0000000000000375

5. Gorelick PB, Scuteri A, Black SE, Decarli C, Greenberg SM, Iadecola C, Launer LJ, Laurent S, Lopez OL, Nyenhuis D, et al. Vascular contributions to cognitive impairment and dementia: a statement for healthcare professionals from the american heart association/american stroke association. Stroke. 2011;42:2672–2713. doi: 10.1161/STR.0b013e3182299496

6. Levine DA, Chen B, Galecki AT, Gross AL, Briceno EM, Whitney RT, Ploutz-Snyder RJ, Giordani BJ, Sussman JB, Burke JF, et al. Associations Between Vascular Risk Factor Levels and Cognitive Decline Among Stroke Survivors. JAMA Netw Open. 2023;6:e2313879. doi: 10.1001/jamanetworkopen.2023.13879

7. Bahader GA, Nash KM, Almarghalani DA, Alhadidi Q, McInerney MF, Shah ZA. Type-I diabetes aggravates post-hemorrhagic stroke cognitive impairment by augmenting oxidative stress and neuroinflammation in mice. Neurochem Int. 2021;149:105151. doi: 10.1016/j.neuint.2021.105151

8. van Sloten TT, Sedaghat S, Carnethon MR, Launer LJ, Stehouwer CDA. Cerebral microvascular complications of type 2 diabetes: stroke, cognitive dysfunction, and depression. Lancet Diabetes Endocrinol. 2020;8:325–336. doi: 10.1016/S2213-8587(19)30405-X

9. Si SC, Yang W, Luo HY, Ma YX, Zhao H, Liu J. Cognitive decline in elderly patients with type 2 diabetes is associated with glycated albumin, ratio of Glycated Albumin to glycated hemoglobin, and concentrations of inflammatory and oxidative stress markers. Heliyon. 2023;9:e22956. doi: 10.1016/j.heliyon.2023.e22956

10. Bartels AL, Leenders KL. Parkinson’s disease: the syndrome, the pathogenesis and pathophysiology. Cortex. 2009;45:915–921. doi: 10.1016/j.cortex.2008.11.010

11. Becker C, Jick SS, Meier CR. Risk of stroke in patients with idiopathic Parkinson disease. Parkinsonism Relat Disord. 2010;16:31–35. doi: 10.1016/j.parkreldis.2009.06.005

12. Skeie GO, Muller B, Haugarvoll K, Larsen JP, Tysnes OB. Parkinson disease: associated disorders in the Norwegian population based incident ParkWest study. Parkinsonism Relat Disord. 2013;19:53–55. doi: 10.1016/j.parkreldis.2012.07.003

13. Huang YP, Chen LS, Yen MF, Fann CY, Chiu YH, Chen HH, Pan SL. Parkinson’s disease is related to an increased risk of ischemic stroke-a population-based propensity score-matched follow-up study. PLoS One. 2013;8:e68314. doi: 10.1371/journal.pone.0068314

14. Struck LK, Rodnitzky RL, Dobson JK. Stroke and its modification in Parkinson’s disease. Stroke. 1990;21:1395–1399. doi: 10.1161/01.str.21.10.1395

15. Marttila RJ, Rinne UK. Arteriosclerosis, heredity, and some previous infections in the etiology of Parkinson’s disease. A case-control study. Clin Neurol Neurosurg. 1976;79:46–56. doi: 10.1016/s0303-8467(76)80005-4

16. Choi HL, Ahn JH, Chang WH, Jung W, Kim BS, Han K, Youn J, Shin DW. Risk of Parkinson disease in stroke patients: A nationwide cohort study in South Korea. Eur J Neurol. 2024;31:e16194. doi: 10.1111/ene.16194

17. Liu Y, Xue L, Zhang Y, Xie A. Association Between Stroke and Parkinson’s Disease: a Meta-analysis. J Mol Neurosci. 2020;70:1169–1176. doi: 10.1007/s12031-020-01524-9

18. Sun F, Salinas AG, Filser S, Blumenstock S, Medina-Luque J, Herms J, Sgobio C. Impact of alpha-synuclein spreading on the nigrostriatal dopaminergic pathway depends on the onset of the pathology. Brain Pathol. 2022;32:e13036. doi: 10.1111/bpa.13036

19. Yan T, Chen Z, Chopp M, Venkat P, Zacharek A, Li W, Shen Y, Wu R, Li L, Landschoot-Ward J, et al. Inflammatory responses mediate brain-heart interaction after ischemic stroke in adult mice. J Cereb Blood Flow Metab. 2020;40:1213–1229. doi: 10.1177/0271678X18813317

20. Feigin VL, Owolabi MO, World Stroke Organization-Lancet Neurology Commission Stroke Collaboration G. Pragmatic solutions to reduce the global burden of stroke: a World Stroke Organization-Lancet Neurology Commission. Lancet Neurol. 2023;22:1160–1206. doi: 10.1016/S1474-4422(23)00277-6

21. Brodtmann A. IJS announces journal series on stroke, cognition and vascular dementia. Int J Stroke. 2011;6:375. doi: 10.1111/j.1747-4949.2011.00662.x

22. Hu G, Jousilahti P, Bidel S, Antikainen R, Tuomilehto J. Type 2 diabetes and the risk of Parkinson’s disease. Diabetes Care. 2007;30:842–847. doi: 10.2337/dc06-2011

23. Cheong JLY, de Pablo-Fernandez E, Foltynie T, Noyce AJ. The Association Between Type 2 Diabetes Mellitus and Parkinson’s Disease. J Parkinsons Dis. 2020;10:775–789. doi: 10.3233/JPD-191900

24. Cerri S, Mus L, Blandini F. Parkinson’s Disease in Women and Men: What’s the Difference? J Parkinsons Dis. 2019;9:501–515. doi: 10.3233/JPD-191683

25. Patel R, Kompoliti K. Sex and Gender Differences in Parkinson’s Disease. Neurol Clin. 2023;41:371–379. doi: 10.1016/j.ncl.2022.12.001

26. Xu Q, Park Y, Huang X, Hollenbeck A, Blair A, Schatzkin A, Chen H. Diabetes and risk of Parkinson’s disease. Diabetes Care. 2011;34:910–915. doi: 10.2337/dc10-1922

27. Lin KD, Yang CY, Lee MY, Ho SC, Liu CK, Shin SJ. Statin therapy prevents the onset of Parkinson disease in patients with diabetes. Ann Neurol. 2016;80:532–540. doi: 10.1002/ana.24751

28. Kawada T. Risk reduction of Parkinson disease by statin therapy in patients with diabetes. Ann Neurol. 2017;81:157. doi: 10.1002/ana.24809

29. Huang X, Alonso A, Guo X, Umbach DM, Lichtenstein ML, Ballantyne CM, Mailman RB, Mosley TH, Chen H. Statins, plasma cholesterol, and risk of Parkinson’s disease: a prospective study. Mov Disord. 2015;30:552–559. doi: 10.1002/mds.26152

30. Huang X, Abbott RD, Petrovitch H, Mailman RB, Ross GW. Low LDL cholesterol and increased risk of Parkinson’s disease: prospective results from Honolulu-Asia Aging Study. Mov Disord. 2008;23:1013–1018. doi: 10.1002/mds.22013

31. Savica R, Grossardt BR, Ahlskog JE, Rocca WA. Metabolic markers or conditions preceding Parkinson’s disease: a case-control study. Mov Disord. 2012;27:974–979. doi: 10.1002/mds.25016

32. Paganini-Hill A. Risk factors for parkinson’s disease: the leisure world cohort study. Neuroepidemiology. 2001;20:118–124. doi: 10.1159/000054770

33. Becker C, Jick SS, Meier CR. Use of antihypertensives and the risk of Parkinson disease. Neurology. 2008;70:1438–1444. doi: 10.1212/01.wnl.0000303818.38960.44

34. Wang YL, Wang YT, Li JF, Zhang YZ, Yin HL, Han B. Body Mass Index and Risk of Parkinson’s Disease: A Dose-Response Meta-Analysis of Prospective Studies. PLoS One. 2015;10:e0131778. doi: 10.1371/journal.pone.0131778

35. Santiago JA, Potashkin JA. System-based approaches to decode the molecular links in Parkinson’s disease and diabetes. Neurobiol Dis. 2014;72 Pt A:84–91. doi: 10.1016/j.nbd.2014.03.019

36. Hernan MA, Takkouche B, Caamano-Isorna F, Gestal-Otero JJ. A meta-analysis of coffee drinking, cigarette smoking, and the risk of Parkinson’s disease. Ann Neurol. 2002;52:276–284. doi: 10.1002/ana.10277

37. Checkoway H, Powers K, Smith-Weller T, Franklin GM, Longstreth WT, Jr., Swanson PD. Parkinson’s disease risks associated with cigarette smoking, alcohol consumption, and caffeine intake. Am J Epidemiol. 2002;155:732–738. doi: 10.1093/aje/155.8.732

38. Reynolds K, Lewis B, Nolen JD, Kinney GL, Sathya B, He J. Alcohol consumption and risk of stroke: a meta-analysis. JAMA. 2003;289:579–588. doi: 10.1001/jama.289.5.579

39. Patra J, Taylor B, Irving H, Roerecke M, Baliunas D, Mohapatra S, Rehm J. Alcohol consumption and the risk of morbidity and mortality for different stroke types--a systematic review and meta-analysis. BMC Public Health. 2010;10:258. doi: 10.1186/1471-2458-10-258

40. Allam MF, Campbell MJ, Hofman A, Del Castillo AS, Fernandez-Crehuet Navajas R. Smoking and Parkinson’s disease: systematic review of prospective studies. Mov Disord. 2004;19:614–621. doi: 10.1002/mds.20029

41. Ritz MF, Ratajczak P, Curin Y, Cam E, Mendelowitsch A, Pinet F, Andriantsitohaina R. Chronic treatment with red wine polyphenol compounds mediates neuroprotection in a rat model of ischemic cerebral stroke. J Nutr. 2008;138:519–525. doi: 10.1093/jn/138.3.519

42. LaHue SC, Comella CL, Tanner CM. The best medicine? The influence of physical activity and inactivity on Parkinson’s disease. Mov Disord. 2016;31:1444–1454. doi: 10.1002/mds.26728

43. Rozani V, Gurevich T, Giladi N, El-Ad B, Tsamir J, Hemo B, Peretz C. Higher serum cholesterol and decreased Parkinson’s disease risk: A statin-free cohort study. Mov Disord. 2018;33:1298–1305. doi: 10.1002/mds.27413

